# Donor-Derived Cell-Free DNA Stratifies Risk of Mortality and Graft Dysfunction in Severe Acute Cardiac Allograft Rejection

**DOI:** 10.1101/2025.09.15.25335831

**Authors:** Zaid N. Safiullah, Han Su, Hyesik Kong, Moon Kyoo Jang, Keyur Shah, Gerald J. Berry, Palak Shah, Hannah A. Valantine, Xin Tian, Sean Agbor-Enoh

## Abstract

**Background:** Cardiac acute rejection (AR) is a risk factor for poor outcomes, however there are limited risk prediction models to stratify patients for death or sustained LV dysfunction. This study assesses the prognostic utility of percentage donor-derived cell-free DNA (%dd-cfDNA) at the diagnosis of AR for poor outcomes.

**Methods:** The prospective multicenter GRAfT study enrolled heart transplant recipients and collected serial plasma samples to quantitate %dd-cfDNA. AR was defined as acute cellular rejection (ACR), antibody-mediated rejection (AMR), as well as biopsy-negative AMR (donor-specific antibody positivity with LV dysfunction). AR was classified as mild-to-moderate (ACR grade 2 or AMR grade 1) or severe (ACR grade ≥3, AMR grade ≥2, or DSA+/LV dysfunction) and further stratified by a %dd-cfDNA threshold of 0.25%. Regression models assessed the association between AR and %dd-cfDNA levels at the AR diagnosis with the primary composite outcome of sustained LVEF decline <50% and/or death.

**Results:** The study included 275 patients and 3,190 %dd-cfDNA assessments. Over the median of 4.6 (IQR 1.8 - 5.0) years follow-up, 51 patients experienced the composite outcome of death or prolonged EF reduction, and 75 patients developed AR, including 16.2% patients with ACR, 9.4% with pathologic AMR, and 6.6% with DSA+/LV dysfunction. Thirty-two (42.7%) patients had severe AR and 43 (57.3%) had mild-to-moderate AR. Severe—but not mild-to-moderate— AR was associated with an increased risk of the primary composite endpoint (HR = 5.17, 95% CI 2.38 − 11.3, p < 0.001). Among those with severe AR, a %dd-cfDNA level greater than 0.25% at diagnosis was associated with a higher risk of the primary outcome (HR, 6.06, 95% CI, 1.78– 20.6; p = 0.004). Percent dd-cfDNA remained elevated in severe AR patients with adverse outcomes.

**Conclusion:** Severe AR with high %dd-cfDNA levels is associated with an increased risk of poor outcomes, offering novel prognostic utility.

**Clinical Perspective:** *What is New?:*

- Percent donor-derived cell-free DNA (%dd-cfDNA) can risk stratify cardiac transplant patients with severe acute rejection for death and/or prolonged EF reduction
- Percent dd-cfDNA remain persistently elevated in patients with severe acute rejection who develop poor outcomes, which could reflect ineffective treatment.
- In the contemporary era of cardiac transplantation, acute rejection defined by biopsy or by donor specific antibodies plus LV dysfunction is associated with poor outcomes

*What are the Clinical Implications?:*

- Percent dd-cfDNA could serve as a bedside tool to risk stratify patients with severe acute rejection for poor outcomes.
- Trends of %dd-cfDNA could serve to monitor response to treatment for severe acute rejection.
- Percent dd-cfDNA levels at diagnosis of rejection could be leveraged for patient selection in clinical trials to test novel therapies or treatment strategies.

## Background

Acute rejection (AR) continues to be a significant cause of morbidity and a risk factor for mortality in the heart transplant recipients despite advances in immunosuppressive therapies^1^. The estimated incidence of AR at two years post-transplant from a large multicenter cohort is 62% ^2^. Acute cellular rejection (ACR) is a common post-transplant complication, with an incidence of 10-14%^3, 4^. The incidence of antibody-mediated rejection (AMR) is variable with a wide range of 3% to 85%, likely due in part to diagnostic limitations^5, 6^. There is currently no available tool in clinical use to identify patients with acute rejection who are at increased risk of poor long-term outcomes.

In the current era of cardiac transplantation, with more effective treatment protocols and updated definitions, the mortality and morbidity associated with acute rejection are not well understood. In prior studies, increasing severity of ISHLT (International Society for Heart and Lung Transplantation) histologic grade in acute rejection and recurrent episodes were associated with increased mortality, development of coronary artery vasculopathy (CAV), and re-transplantation^7–9^. These sometimes decades-old studies that established these associations were often single-center, retrospective and/or did not use more contemporary definitions of AMR or include EMB-negative forms of AMR^10–12^, which are important risk factors for mortality^13^ and CAV ^14, 15^. The studies also rely on EMB for grading AR severity, an invasive approach limited by a high interrater variability to grade acute rejection and a low diagnostic yield, even when patients present with significant allograft dysfunction ^16^. Noninvasive biomarkers, such as percent donor-derived cell-free DNA (%dd-cfDNA) now presents sensitive alternatives to assess allograft injury and severity^1, 17, 18^ . Elevated levels of %dd-cfDNA are associated with both contemporarily defined ACR, AMR, and subclinical injury, often preceding histological changes detectable by EMB ^18^, as well as with EMB-negative AMR^13^. Percent dd-cfDNA could, therefore, capture severe forms of AR associated with important clinical outcomes.

The lack of reliable clinical endpoints also limits risk stratification in heart transplantation. While mortality remains the most definitive adverse outcome, the low incidence limits clinical trial design. Significant and sustained reductions in left ventricular (LV) function represent a critical intermediate endpoint that strongly correlates with morbidity, quality of life, and long-term graft survival, even in the absence of high-grade histologic rejection ^12^. Historically, both ACR and AMR are associated with echocardiographic allograft dysfunction^10, 11, 19^. A composite endpoint of sustained reduction in LVEF and/or mortality could therefore provide intermediate and long-term benchmark to assess the risk of acute rejection.

This multicenter, prospective cohort study aims to evaluate the prognostic significance of %dd-cfDNA levels measured at the time of acute rejection diagnosis and to investigate the association between acute rejection and mortality and morbidity. We hypothesize that elevated %dd-cfDNA levels are associated with an increased risk of mortality and prolonged reduction in LVEF., and we posit that acute rejection is associated with mortality and sustained allograft dysfunction even in the current era with advances in treatment strategies. By elucidating the prognostic utility of %dd-cfDNA, our goal is to enhance risk stratification and guide personalized management strategies for patients with acute rejection.

## Methods

### Study Design

The GRAfT (Genomic Research Alliance for Transplantation Study, NCT02423070) study enrolled heart transplant recipients aged 18 years or older on the waitlist before the transplant and followed them serially after the transplant. Patients undergoing repeat or multi-organ heart transplantation were excluded. Patients were recruited at five regional transplant centers: Inova Schar Heart and Vascular, The Johns Hopkins Hospital, the University of Maryland Medical Center, Virginia Commonwealth University, and MedStar Washington Hospital Center. The full study methodology, including center immunosuppression and surveillance protocols, has been previously published ^18^. We collected data between 2015 and 2025. The institutional review boards of all centers and the NHLBI approved the study, and patients provided their informed consent before enrollment. This study adheres to the principles of the Declaration of Helsinki and the ISHLT statement on Transplant Ethics. The data that support the findings of this study are available from the corresponding author upon reasonable request.

### Post-transplant Evaluations

Patients underwent surveillance and clinically indicated EMB and other evaluations, as part of routine clinical care. The individual center’s pathologist used the ISHLT grading system to grade for AMR and ACR. We defined AMR as pAMR 1 (H+), 1 (I+), 2, or 3. We also include a recent category synonymous to EMB-negative AMR defined as a positive DSA evaluation with LV dysfunction, defined as an LVEF decrease by ≥10% from the previous measurement to an LVEF ≤50%^13^. Luminex (Luminex, Austin, TX) multiplex bead assays were used to evaluate the presence, phenotype, and quantity of DSA, with an MFI threshold of 1000 used to determine a positive DSA evaluation. We defined mild-to-moderate acute rejection as ACR grade 2 or pAMR grade 1, and severe acute rejection, defined as ACR grade ≥3, pAMR grade ≥2, or DSA+LV dysfunction. We defined control subjects as those without pAMR+, ACR+, or DSA-positive findings with left ventricular (LV) dysfunction.

### Outcome Measures

The primary composite endpoint was death and/or prolonged EF reduction (≤50%) for ≥90 days). We excluded death events occurring within the first six months of the transplant. The secondary outcome was death occurring by the end of the follow-up period. Prolonged EF reduction was not used as an outcome measure due to a paucity of events.

### Percent dd-cfDNA Measurement

We genotyped the donor and recipient DNA to identify informative single-nucleotide polymorphisms. After transplantation, we isolated plasma cfDNA for library construction and paired-end shotgun sequencing. Sequence reads were analyzed to identify donor, and recipient reads using informative single-nucleotide polymorphisms. We computed %dd-cfDNA as a percentage of reads with donor single-nucleotide polymorphisms to reads of donor plus recipient single-nucleotide polymorphisms ^18, 20^. Given known early post-transplant exponential decay^21^, dd-cfDNA measurements <28 days post-transplant were excluded. Elevated %dd-cfDNA was defined as ≥0.25%, as measurement by a previous study ^18^.

### Statistical Methods

Descriptive statistics were used to summarize baseline recipient and transplant characteristics. Continuous variables are reported as medians (interquartile range [IQR]), and categorical variables as counts with percentages. Cumulative incidences for pAMR, ACR, or DSA+/LV dysfunction, and their composite were estimated, treating earlier death without AR as a competing risk. Kaplan–Meier curves described the probabilities of survival and the composite outcome of prolonged EF reduction and death. For each patient, the AR episodes and outcomes occurring ≥30 days after transplant were retained, excluding peri-operative events (<30 days). Levels of %dd-cfDNA at the diagnosis of AR were log10-transformed and compared to the control patients without AR (AR-). Every AR case was time-matched (± 1 day) 1:2 to AR- controls. Differences in the mean %dd-cfDNA were assessed using a linear mixed model that included patient subgroup as a fixed effect and a subject-specific random intercept to account for repeated measures. To examine the association with prolonged EF reduction and death, univariate and multivariable Cox regression analyses were performed, with the occurrence of the earliest AR as a time-dependent covariate. Furthermore, we assessed the association of transplant outcomes with AR by different severity and AR by severity with further dichotomization using %dd-cfDNA at diagnosis > 0.25. Hazard ratios (HRs) and 95% confidence intervals (CIs) were reported. All tests were two-sided with P < 0.05 denoting statistical significance. Statistical analyses were conducted using R statistical software (version 4.5.0, R Foundation for Statistical Computing).

## Results

### Patient Population

The study enrolled 281 heart transplant recipients between July 2015 and September 2020. After excluding six patients who died within six months of transplant or had insufficient clinical data, 275 patients were included in the analysis (Figure 1). The median age was 56 years (IQR 48-63); 74.4% were men, 48.7% were White, and 58.5% of patients underwent transplantation for a nonischemic cardiomyopathy. A summary of demographic characteristics is represented in Table 1. Patients with and without acute rejection showed similar demographic and baseline characteristics (Table S1).

**Figure 1.**
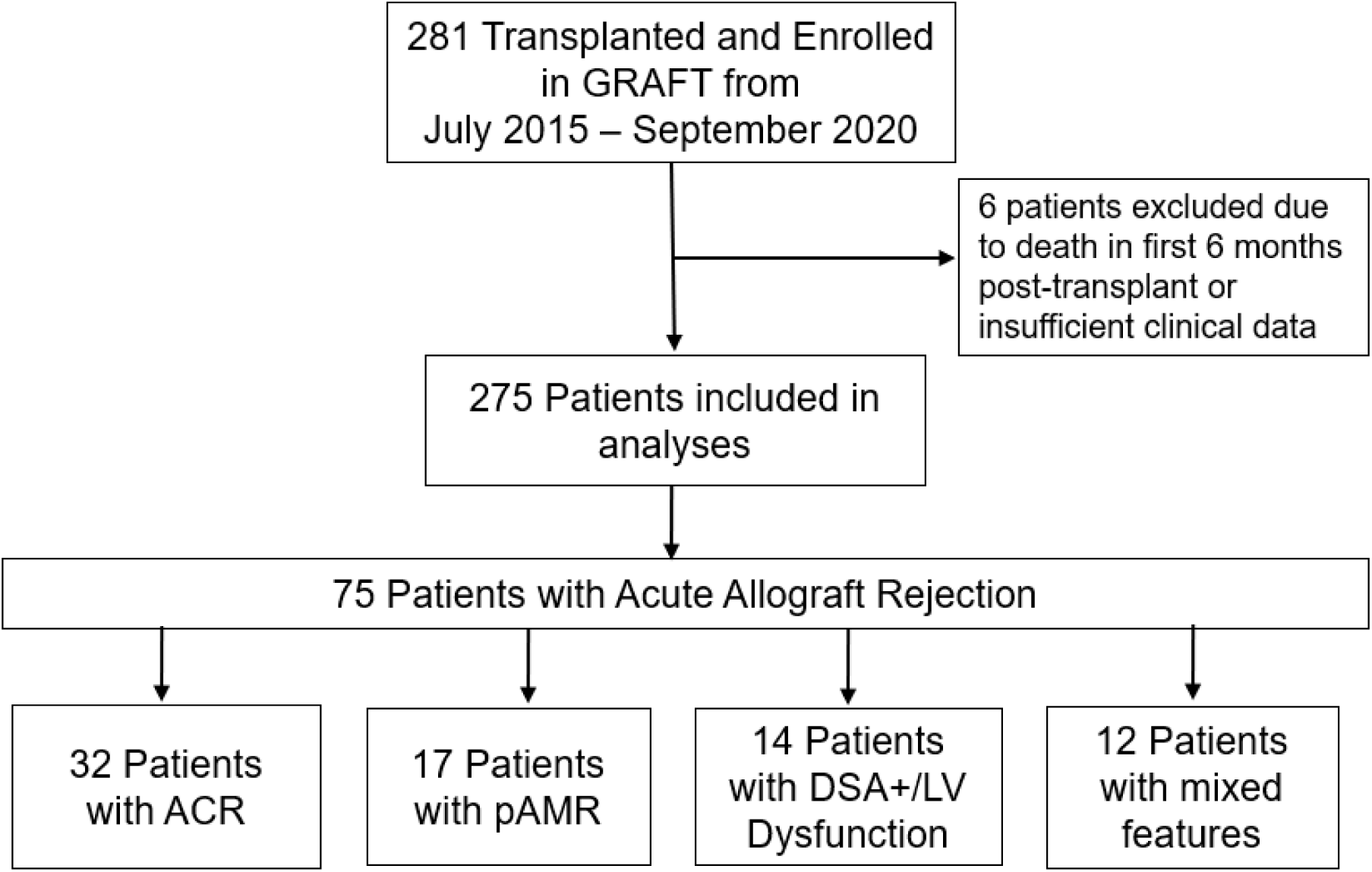
Flow diagram of study participants

**Table 1.** Baseline Patient and Transplant Characteristics.

| Characteristic | Overall (N=275) |
| --- | --- |
| **Age, yr** |  |
| median (IQR) | 56 (48, 63) |
| **Sex** |  |
| Women | 70 (25.5%) |
| Men | 205 (74.5%) |
| **Race** |  |
| Black | 114 (41.5%) |
| White | 130 (47.3%) |
| Hispanic | 20 (7.3%) |
| Asian | 6 (2.2%) |
| Multiracial | 5 (1.8%) |
| **Ethnicity** |  |
| Latino or Hispanic | 21 (7.6%) |
| Not Latino or Hispanic | 254 (92.4%) |
| **Smoking history** |  |
| No | 163 (59.3%) |
| Yes | 107 (38.9%) |
| Unknown | 5 (1.8%) |
| **BMI** |  |
| <30 | 175 (63.6%) |
| ≥30 | 94 (34.2%) |
| Unknown | 6 (2.2%) |
| **Primary diagnosis** |  |
| Ischemic cardiomyopathy | 76 (27.6%) |
| Nonischemic cardiomyopathy | 157 (57.1%) |
| Hypertrophic cardiomyopathy | 8 (2.9%) |
| Myocarditis | 6 (2.2%) |
| Restrictive cardiomyopathy | 8 (2.9%) |
| Other | 20 (7.3%) |
| **VAD usage** |  |
| No | 183 (66.5%) |
| Yes | 82 (29.8%) |
| Not Reported | 10 (3.6%) |
| **UNOS status** |  |
| Status 1A | 139 (50.5%) |
| Status 1B | 57 (20.7%) |
| Other | 79 (28.7%) |

*Note:* Data are n(%) or median (IQR),

Figure 2a shows the cumulative incidence of acute rejection categories. Over a median follow-up of 4.6 years (IQR 1.8 – 5.0), 26.9% of patients developed AR, including 16.2% with ACR, 9.4% with pathologic AMR, and 6.6% with DSA+/LV dysfunction. The breakdown of patients with different types of AR is shown in Figure 1. Thirty-two patients (11.6%) experienced severe AR (64 episodes), while 43 patients (15.6%) had only mild-to-moderate acute rejection (79 episodes).

**Figure 2.**
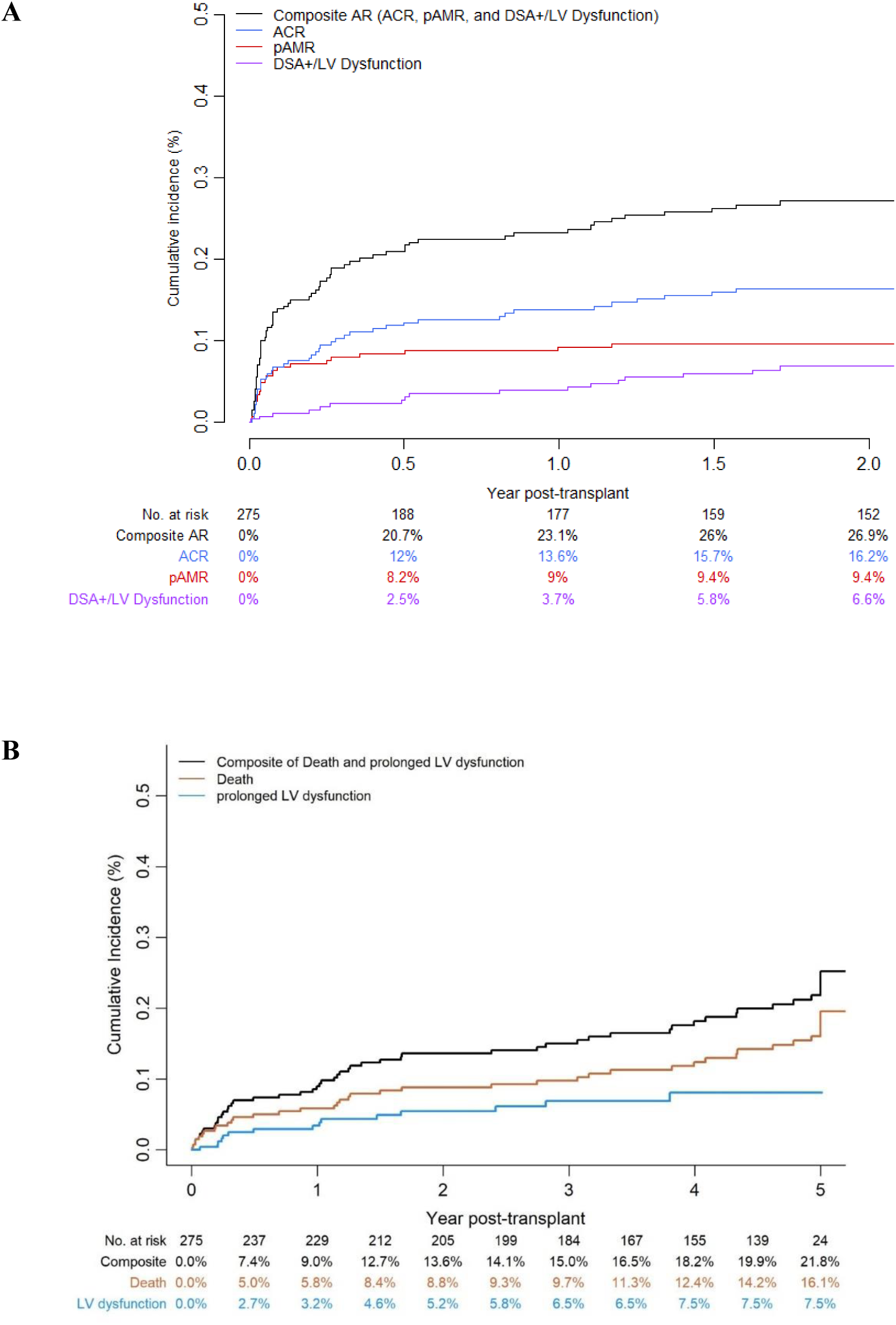
A) Cumulative Incidence of Acute Rejection, including ACR, pAMR, AMR by DSA+/LV dysfunction and the composite of these events. B) Cumulative Incidence of Outcomes. The composite outcome includes the prolonged EF reduction or death.

The cumulative incidence of the primary composite endpoint and secondary endpoints is shown in Figure 2b. At the 5-year follow-up, 51 patients died with an estimate survival rate of 83.9%; 21.8% experienced the composite outcome of death (16.1%) or prolonged EF reduction (7.5%), while 78.2% were alive without prolonged EF reduction.

### Acute Rejection is Associated with Poor Outcomes

Acute rejection was associated with an increased risk of the composite primary endpoint of death and sustained EF reduction in univariate (HR 4.23, 95% CI, 2.25 − 7.94, p < 0.001) and multivariable models that adjusted for age, sex, and race (adjusted HR 4.12, 95% CI 2.19 − 7.75, p < 0.001, Table 2). In multivariable analysis, the risk remained significant for the different phenotypes of acute rejection, AMR defined by biopsy or DSA+/LV dysfunction (HR 2.91, 95% CI 1.37 − 6.20, p = 0.006), and ACR (HR 4.88, 95% CI 2.37 − 10.0, p < 0.001) (Table 2). Acute rejection, as well as its phenotypes were also associated with an increased risk of death (Table 2).

**Table 2.** Association of Acute Rejection with the Risk of Death and Prolonged EF Reduction.

|  | Composite outcome<br>(prolonged EF Reduction /death) |  | Death |  |
| --- | --- | --- | --- | --- |
|  | HR (95% CI) | *P* | HR (95% CI) | *P* |
| Unadjusted models |  |  |  |  |
| AR | 4.23 (2.25, 7.94) | <0.001 | 3.18 (1.48, 6.84) | 0.003 |
| AMR | 3.14 (1.48, 6.64) | 0.003 | 3.12 (1.31, 7.41) | 0.01 |
| ACR | 4.46 (2.21, 8.98) | <0.001 | 2.76 (1.11, 6.84) | 0.029 |
| Multivariable models |  |  |  |  |
| AR | 4.12 (2.19, 7.75) | <0.001 | 2.93 (1.35, 6.33) | 0.006 |
| AMR | 2.91 (1.37, 6.20) | 0.006 | 2.69 (1.12, 6.47) | 0.027 |
| ACR | 4.88 (2.37, 10.0) | <0.001 | 2.72 (1.08, 6.83) | 0.034 |

*Note:* Acute Rejection (AR) is defined as the composite event of acute cellular rejection (ACR), antibody-mediated rejection (AMR) determined by EMB or DSA+ with LV dysfunction. Cox regression models were used with a time-dependent covariate for the development of AR, AMR or ACR. Multivariable analysis was adjusted for patient's age, sex, race.

When AR as stratified by severity, i.e. severe or mild-to-moderate, and adjusted for patient’s age, sex, and race, severe acute rejection was associated with risk of the composite outcome (HR 5.17, 95% CI 2.38−11.3, p < 0.001) and death (HR 4.76, 95% CI 1.90−11.9, p < 0.001, Table 3).

**Table 3:** Multivariable analysis of AR by severity and %dd-cfDNA levels with risk of poor outcomes.

|  | Composite outcome<br>(prolonged EF Reduction /death) |  | Death |  |
| --- | --- | --- | --- | --- |
|  | HR (95% CI) | p | HR (95% CI) | p |
| **AR severity** |  |  |  |  |
| Mild-to-moderate | 2.37<br>(0.90, 6.25) | 0.081 | 2.31<br>(0.77, 6.89) | 0.13 |
| Severe | 5.17<br>(2.38, 11.3) | <0.001 | 4.76<br>(1.90, 11.9) | <0.001 |
| **AR severity and %dd-cfDNA categorization** |  |  |  |  |
| Mild-to-moderate with<br>ddcfDNA<.25% | 2.55<br>(0.87, 7.52) | 0.09 | 2.26<br>(0.65, 7.81) | 0.20 |
| Mild-to-moderate with<br>ddcfDNA≥.25% | 1.98<br>(0.27, 14.6) | 0.51 | 2.51<br>(0.33, 18.9) | 0.37 |
| Severe with ddcfDNA<.25% | 1.57<br>(0.37, 6.66) | 0.54 | 0.72<br>(0.10, 5.50) | 0.75 |
| Severe with ddcfDNA≥.25% | 6.06<br>(1.78, 20.6) | 0.004 | 10.3<br>(2.85, 37.3) | <0.001 |

*Note:* Cox regression models were used with a time-dependent covariate for the AR by grade only or AR by grade and %dd-cfDNA at diagnosis. Multivariable analysis was adjusted for patient's age, sex, race. AR severity was determined as mild-to-moderate (ACR grade 2 or pAMR grade 1) and severe (ACR grade ≥3, pAMR grade ≥2, or DSA+/LV dysfunction [donor-specific antibody presence and LVEF drop ≥10% to a value ≤50%]).

Severe AR defined by biopsy alone also showed increased risk of poor outcomes in multivariable and univariate models (Table S2). However, mild-to-moderate AR showed a non-significant trend associated with the composite outcome and death, respectively. The unadjusted univariate models of AR severity showed similar results (Table S3).

### Percent dd-cfDNA is associated with Poor Clinical Outcomes in Acute Rejection

Consistent with prior reports, %dd-cfDNA values were elevated for acute rejection, and its phenotypes compared to no rejection controls (Figure 3, Figure S1). Severe AR showed approximately 2-fold higher than mild-to-moderate AR (0.38% vs 0.19%, p =0.002).

**Figure 3.**
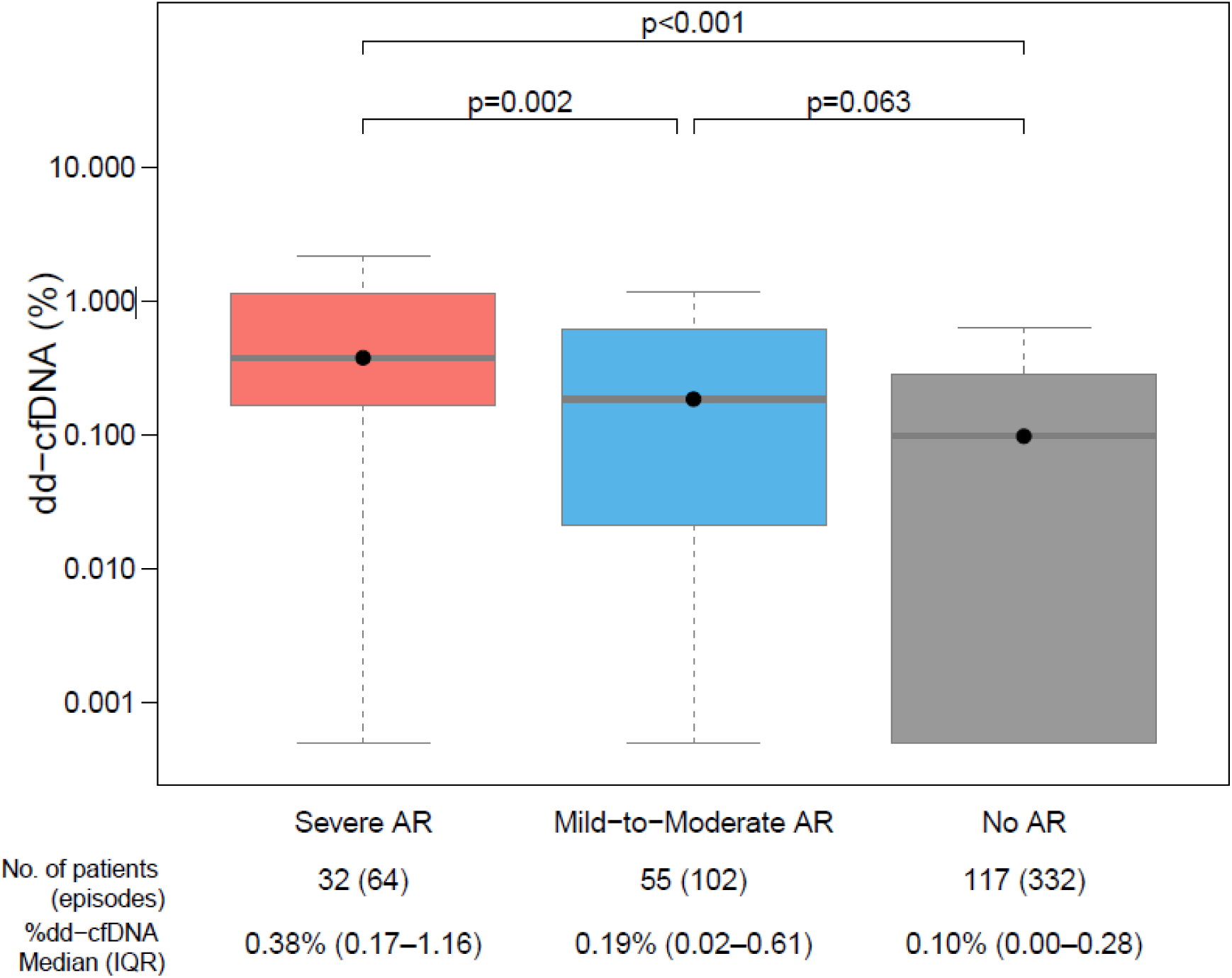
Donor-derived cell-free DNA (%) at the time of AR diagnosis for different AR groups and at matched times for controls with no AR. Severe AR (*ACR grade ≥3, pAMR grade ≥2, or DSA+/LV dysfunction*); mild-to-moderate AR (ACR grade 2 or pAMR grade 1) and controls with no AR, with %dd-cfDNA measured at 2:1 time-matched to the time of AR diagnosis.

Within the acute rejection categories, risk differed based on %dd-cfDNA levels (Table 3). In multivariable analyses, severe AR with %dd-cfDNA ≥ 0.25% was associated with an increased risk of prolonged EF reduction /death (HR 6.06, 95% CI 1.78 − 20.6, p = 0.004) and death (HR 10.3, 95% CI 2.85 − 37.3, p < 0.001). This was consistent with the univariable analysis (Table S3). Severe AR defined by biopsy alone and %dd-cfDNA ≥ 0.25% showed similar increased risk for poor outcomes in multivariable and univariate models (Table S4). However, severe AR with %dd-cfDNA < 0.25% was not associated with increased risk of the composite outcome (HR 1.57 95% CI 0.37- 6.66, p=0.54), nor death (HR 0.72 95% CI 0.10- 5.50, p=0.75), Mild-to-moderate acute rejection with and without %dd-cfDNA ≥ 0.25% was not associated with an increased risk of death or the composite of death or sustained reduction in LVEF (Table 3).

The univariable analyses were consistent (Table S3).

### Percent dd-cfDNA trends over time

Treatment received for acute rejection is summarized in Table S5. The post-treatment and post- AR %dd-cfDNA trends differed based on the subsequent development of the primary outcome; patients who died or showed sustained LV reduction exhibited higher %dd-cfDNA at AR diagnosis and persistent elevation over the post-transplant period above baseline levels. On the contrary, the patients who did not develop these adverse outcomes showed decreasing levels over time; the levels approached the pre-rejection baseline (Figure S2).

## Discussion

Despite advances in treatment, acute rejection remains a leading cause of morbidity and mortality following cardiac transplantation^1, 7–9^. Unfortunately, risk stratification remains challenging. In this large multicenter prospective cohort, we demonstrated that severe and not mild-to-moderate acute rejection is associated with death and a composite outcome of death or sustained reduction in LVEF. In the severe acute rejection category, elevated %dd-cfDNA >0.25% at diagnosis was associated with an increased risk. The patients who go on to develop adverse outcomes show persistently elevated %dd-cfDNA at and following diagnosis and despite treatment for acute rejection. We also demonstrated that in the contemporary era of cardiac transplantation, acute rejection, defined by current ISHLT histologic grading^1^, and EMB- negative AMR additionally defined by DSA+/ LV dysfunction^13^, are associated with death and the composite outcome. Our study established potential %dd-cfDNA thresholds that could be utilized at the bedside to risk stratify patients with severe acute rejection for poor outcomes.

Patients who develop severe acute rejection (ACR grade 3 or pAMR 2) received similar therapies. However, they exhibit different allograft injury profiles at diagnosis and after treatment. Those who had %dd-cfDNA levels above a prespecified threshold of 0.25% were at significantly higher risk for adverse outcomes. The prespecified %dd-cfDNA threshold was shown to be predictive of poor outcomes. These high-risk patients also showed persistent elevations in %dd-cfDNA after diagnosis and treatment, whereas the low-risk patients showed decreased %dd-cfDNA trends toward the pre-rejection baseline. The persistently elevated %dd-cfDNA could indicate an inadequate treatment response and a need for additional or different treatment strategies. These patients could benefit from second-line therapies or more prolonged treatment duration. The %dd-cfDNA patterns could also be used to select candidates for clinical trials to test new treatment strategies. A biomarker-driven treatment strategy could improve outcomes by targeting therapy to those most at risk while minimizing unnecessary immunosuppression and its associated complications. Notably, we did not observe similar findings in those with mild-to-moderate acute rejection (ACR grade 2 or pAMR 1), who also received treatment for acute rejection. The finding could have been limited by the small sample size of patients with mild-to-moderate acute rejection, successful treatment responses that may have derisked these patients for downstream outcomes, or other unidentified reasons. The %dd-cfDNA trends over the post-transplant period in those with severe acute rejection or functional AMR may offer mechanistic insights into ongoing allograft injury.

The contemporary era in cardiac transplant is marked by rigorous monitoring for early detection and treatment of acute rejection to derisk patients for unfavorable outcomes. This study utilizes %dd-cfDNA as a sensitive, noninvasive biomarker that is commercially available, which is a departure from prior pioneering studies that established the risk association of AR and death using EMB. Assessments of %ddcfDNA are reproducible, not liable to significant measurement variability ^20^, and sensitive to the different categories of acute rejection, including the recently described category of potentially EMB-negative AMR^13^. Gene expression profiling (GEP) is an alternative noninvasive approach used for surveillance in low-risk transplant recipients. It has not been studied for risk assessments in patients with biopsy-confirmed rejection or biopsy-negative forms of rejection ^22–24^. In addition to the use of %dd-cfDNA, this study assessed the association of AR to an intermediate outcome, which is increasingly recognized as a potential harbinger of early death ^7–9^. The study also includes additional forms of rejection, which were not previously assessed.

Our study has several limitations. The sample size was modest, with a small number of patients experiencing acute rejection, particularly in the DSA and LV dysfunction categories. The prevalence of the composite outcome was also low. These event rates may have limited our power to detect an association between %dd-cfDNA and outcomes in patients with mild-to-moderate acute rejection. The %ddcfDNA threshold proposed has been reported in prior studies ^13, 18^ but differs from the thresholds used in other cohort studies ^25, 26^. The study did not evaluate the effect of treatment on the effectiveness of treating acute rejection and on the risk of poor outcomes. Future studies with larger sample sizes may validate these %dd-cfDNA thresholds and address the other limitations. Future studies should assess whether prolonged treatment in high-risk patients or other approaches could reduce the risk of these adverse outcomes. Future studies should also assess the reversibility of the intermediate outcome used in this study and assess the generalizability of these findings in other cohorts. Our findings should be viewed as hypothesis-generating, pending further assessments in future studies.

In conclusion, in this contemporary era with new definitions and rigorous patient monitoring, acute rejection remains a risk factor for both death and an intermediate endpoint. Our findings suggest that risk remains significant in patients with severe forms of acute rejection, particularly in those with elevated %dd-cfDNA at diagnosis. These patients showed persistently elevated %dd-cfDNA after treatment. Our findings suggest that %dd-cfDNA could be a promising noninvasive biomarker, enabling the monitoring of treatment response and risk stratification for mortality. Pending validation studies, these findings support the integration of %dd-cfDNA into clinical risk decision-making in heart transplant recipients.

## Data Availability

The data that support the findings of this study are available from the corresponding author upon reasonable request.

## Sources of Funding

This research was supported in part by the Intramural Research Program of the National Institutes of Health (NIH). The contributions of the NIH authors were made as part of their official duties as NIH federal employees, are in compliance with agency policy requirements, and are considered Works of the United States Government. However, the findings and conclusions presented in this paper are those of the authors and do not necessarily reflect the views of the NIH or the U.S. Department of Health and Human Services.

## Disclosures

The authors have declared that no conflict of interest exists.

## Supplemental Material

Tables S1–S5

Figures S1 and S2

## Notes

### Competing Interest Statement

The authors have declared no competing interest.

### Clinical Trial

Genomic Research Alliance for Transplantation Study, NCT02423070, ClinicalTrials.gov

### Author Declarations

The institutional review boards of all centers and the NHLBI approved the study, and patients provided their informed consent before enrollment. This study adheres to the principles of the Declaration of Helsinki and the ISHLT statement on Transplant Ethics.

